# Impact of opioid overdoses on US life expectancy and years of life lost, by demographic group and stimulant co-involvement: a mortality data analysis from 2019-2022

**DOI:** 10.1101/2023.07.07.23292297

**Authors:** Anne H. Hébert, Alison L. Hill

## Abstract

**Background:** The United States’ opioid crisis is worsening, and the number of deaths reached 81,806 in 2022 after more than tripling over the past decade. This study aimed to comprehensively characterize changes in burden of opioid overdose mortality in terms of life expectancy reduction and years of life lost between 2019-2022, including differential burden across demographic groups and the contribution of polysubstance use.

**Methods:** Using life tables and counts for all-cause and opioid overdose deaths from the National Center for Health Statistics, we constructed cause-eliminated life tables to estimate mortality by age in the absence of opioid-related deaths. We calculated the loss in life expectancy at birth (LLE) and total years of life lost (YLL) due to opioid overdose deaths by state of residency, sex, racial/ethnic group, and co-involvement of cocaine and psychostimulants.

**Findings:** Opioid-related deaths in the US led to an estimated 3·1 million years of life lost in 2022 (38 years per death), compared to 2·0 million years lost in 2019. Relative to a scenario with no opioid mortality, we estimate that opioid-related deaths reduced life expectancy nationally by 0·67 years in 2022 vs 0·52 years in 2019. This LLE worsened in all racial/ethnic groups during the study period: 0·76y to 0·96y for white men, 0·36y to 0·55y for white women, 0·59y to 1·1y for Black men, 0·27y to 0·53y for Black women, 0·31y to 0·82y for Hispanic men, 0·19y to 0·31y for Hispanic women, 0·62y to 1·5y for American Indian/Alaska Native (AI/AN) men, 0·43y to 1y for AI/AN women, 0·09y to 0·2y for Asian men, and 0.08y to 0.13y for Asian women. Nearly all states experienced an increase in years of life lost (YLL) per capita from 2019-2022, with YLL more than doubling in 16 states. Cocaine or psychostimulants with abuse potential (incl. methamphetamines) were involved in half of all deaths and years of life lost in 2022, with substantial variation in the predominant drug class by state and racial/ethnic group.

**Interpretation:** The burden of opioid-related mortality increased dramatically in the US between 2019-2022, coinciding with the period of the COVID-19 pandemic and the associated disruptions to social, economic, and health systems. Opioid overdose deaths are an important contributor to decreasing US life expectancy, and Black, Hispanic, and Native Americans now experience mortality burdens approaching or exceeding white Americans.

**Funding:** None

## Research in context

### Evidence before this study

The opioid crisis in the US is increasing in severity, with more opioid overdose deaths occurring each year. We searched PubMed and Google Scholar with the terms “opioid epidemic”, “opioid crisis”, “opioid overdoses”, or “opioid mortality” to identify studies evaluating the progression of opioid mortality around the world, and in particular in the US. We used the additional terms “stimulants”, “psychostimulants” or “polysubstance use” to find studies of the increasing polysubstance use, and we used the additional terms “demographic”, or “race and ethnicity” to find studies of the shifting demographics. We used the additional terms “life expectancy” or “life years lost” to find studies estimating these metrics of burden. When finding relevant papers through these searches, we also looked at their citations, as well as the authors’ other publications.

Previous work has established that opioid mortality is a rapidly changing public health crisis in the US. The years of life lost to opioid overdose deaths have been estimated at various stages of this crisis and for various subpopulations. One study estimated 1.7 million years of life lost to opioid overdoses in the US in 2016, compared to more recent work finding 2.9 million years lost to unintentional opioid overdoses alone in 2021. Another study estimated that the US population lost 0·36 years in life expectancy as a result of opioid overdose deaths in 2016. However, we could not find estimates of the impact of opioid overdose deaths on US life expectancy or years of life lost in recent years, or comprehensive comparisons of these metrics across demographic groups. We also could not find studies estimating the contributions of the increasing co-involvement of stimulants and opioids to these metrics.

### Added value of this study

We used all-cause and opioid-related mortality counts to estimate the impact of opioid overdose deaths on life expectancy at birth each year from 2019-2022, using cause-eliminated life tables. We found that opioid-related mortality in 2022 reduced US life expectancy by 0·67 years (compared to 0·52 years in 2019), and resulted in the loss of 3·1 million years of life (compared to 2 million years lost in 2019) or 38 life years lost per individual death (39 in 2019). We estimated these metrics of burden comprehensively across demographic groups, finding up to 10-fold variation by race/ethnicity and sex or by state. We observed increases in nearly all groups from 2019-2022; nationally, years of life lost increased from 34% among white women to 170% among American Indian/Alaska Native men. The loss in life expectancy at birth was worst amongst AI/AN and Black men, at 1·5 and 1·1 years, respectively. We also quantified the contribution of polysubstance use to these metrics of burden, finding that in 2022 polysubstance use of opioids in combination with stimulants was responsible for half the years of life lost to opioid-related deaths.

### Implications of all the available evidence

Our findings build on prior work to highlight the substantial contribution of opioid-related deaths to the decrease in life expectancy in the US. The young age at which opioid overdose deaths occur - compared to other leading causes of death in the US - amplifies the impact of this crisis on overall life expectancy and life years lost. Our results and previous evidence confirm that the demographics of the opioid crisis have shifted in recent years and that the mortality burden of opioid overdoses is now increasing across all demographic groups in the US. Additionally, the growing co-involvement of stimulants such as methamphetamine and cocaine with opioids is leading to a considerable loss of life. Understanding the mortality burden and the populations at highest risk is crucial for effectively implementing and delivering treatments.

Despite substantial federal and local investments to slow the opioid crisis over the past decade, including expansions to opioid-use-disorder treatments included in COVID-19 response and recovery funding, opioid overdose mortality has only accelerated in recent years. Transformative policy approaches to the opioid crisis are urgently needed to overturn the trends of decreasing life expectancy and to avoid the annual loss of millions of years of life.

## Introduction

The opioid epidemic is an urgent public health issue in the United States. Fatal opioid overdoses in the US have increased more than tenfold since 1999, reaching 81,806 deaths in 2022.^1^ The initial rise in opioid-related deaths was driven by prescription opioids, following aggressive marketing by the pharmaceutical industry.^2,3^ A second wave, consisting of increasing heroin overdose deaths emerged in 2010, followed in 2013 by a third wave involving synthetic opioids (primarily illicitly manufactured fentanyl). Most recently, a fourth wave of polysubstance use has emerged, in which stimulants such as methamphetamines or cocaine are increasingly used in combination with opioids.^4^ This increased co-use of stimulants and opioids, associated with a higher risk of fatal overdose, is hypothesized to be driven by a combination of users’ desire for the combined effects of the substances and their pervasiveness in the drug supply.^5,6^ Following the onset of the COVID-19 pandemic, fatal and non-fatal overdoses involving opioids and combinations of opioids and stimulants surged.^4,7^ While in its initial waves the opioid crisis was portrayed as predominantly affecting white people, the demographics have shifted in recent years.^8^ Disproportionately affected by the fourth wave of polysubstance use, Black/African American and American Indian/Alaska Native people are now experiencing higher opioid-related mortality rates than white people.^9^

An estimated 7 million people in the US had opioid use disorder (OUD) in 2019.^10^ OUD includes addiction, increased tolerance, and withdrawal symptoms when use is stopped, and contributes to health and social problems. A study estimated that individuals with OUD had 10 times higher mortality than the general population, and experienced overdose death rates of about 8 per 1,000 person-years, in the years 2006-2014.^11^ While effective medications exist for OUD, it is estimated that only 28% of people needing treatment receive it.^12^ Treatment initiation and retention rates are even lower among racial and ethnic minorities, due to disparities in access and systemic biases in health care.^8^

Opioid overdose mortality rates are alarmingly high, but measures based on death counts alone do not fully capture the impact of this crisis as they do not account for the young age at which most opioid-related deaths occur, compared to other leading causes of death like heart disease, stroke, cancer, and COVID-19. Consequently, calculating the impact of opioid mortality on US life expectancy better accounts for the age distribution of deaths. Similarly, years of life lost (YLL) estimates how many additional years individuals would have lived, had they not died prematurely from the cause in question.^13^ Previous studies have estimated the YLL to the opioid crisis, both nationwide and for various subpopulations,^14–18^ and examined past impacts of opioid-related deaths on the US life expectancy at birth.^19^ However, the impact of this recent surge in opioid-related deaths on the US life expectancy has not been estimated. There are also no updated, comprehensive comparisons of YLL across race and ethnicity groups since the observed shifts in the demographics of the crisis and the onset of COVID-19, nor have the YLL to stimulant co-involvement been estimated.

In this study, we quantify the changes in opioid-related mortality in the years 2019, 2020, 2021, and 2022, coinciding with the onset of the COVID-19 pandemic. For each year, we estimate the reduction in the US population’s life expectancy at birth due to opioid overdose deaths, in addition to estimating the resulting years of life lost. We compare these metrics across demographic groups, highlighting which groups have seen the largest loss in life expectancy due to opioid-related mortality in the last few years. We also estimate the contributions of polysubstance use, both nationwide and by demographic group.

## Methods

### Data

This study uses data from the National Center for Health Statistics’s Multiple Cause of Death database, accessible from CDC WONDER (Wide-ranging ONline Database for Epidemiological Research). For each study year (2019, 2020, 2021, and 2022), we accessed the number of deaths occurring that year, and crude and age-adjusted death rates, for all-cause and opioid-related deaths, stratifying by age, sex, race, Hispanic origin, and/or state of residency. At the time of writing, 2022 was the most recent year for which official death counts were available. Provisional death counts may be incomplete due to reporting delays that vary by cause of death and jurisdiction and disproportionately affect overdose deaths. Age-adjusted mortality rates reported here used the 2000 Standard Population. The mortality data are based on death certificates of US residents in all counties. Causes of death are categorized by Underlying Cause of Death (UCD) and by Multiple Cause of Death (MCD), using ICD-10 codes. For opioid overdose deaths, we filter for deaths with a UCD code indicating drug poisoning (X40-44: unintentional, X60-64: suicide, X85: homicide, Y10-14: undetermined intent) and an MCD code indicating an opioid (T40.0-T40.4: Opium, Heroin, Natural and Semisynthetic Opioids, Methadone, and Synthetic Opioids, and T40.6: Other and Unspecified Narcotics).^2^ Overdose deaths involving combinations of opioids and stimulants are identified by opioid MCD codes in addition to the stimulant’s MCD code (T40.5: cocaine, T43.6: psychostimulants with abuse potential).^9^ Any opioid-related deaths involving both additional substances would appear in both categories since WONDER does not allow for additional filtering. However, we can also filter death counts to include all those that involved either psychostimulants or cocaine or both, thus avoiding this double counting when estimating metrics of total stimulant co-involvement.

CDC WONDER reports sex in 2 categories: male or female (refer to *Supplementary Methods* for details). We used the option to subdivide into 6 race categories, with Hispanic ethnicity reported separately. Race and Hispanic ethnicity are determined by the funeral director based on information given (e.g. a surviving next of kin) or by observation.^1^ We combine race and ethnic origin, forming 7 groups: one containing all individuals of Hispanic or Latino origin and of any race (96% identified as white), and 6 groups containing individuals of Non-Hispanic or Latino origin, grouped by race (**Table 1**). We did not further analyze the ‘More than one race’ or NHOPI groups, or individuals with “Not Stated” Hispanic origin when stratifying by race and ethnicity since there are no NVSS life tables for the former group, and latter two groups have very few opioid-related deaths.

**Table 1:** Opioid-related mortality rates in 2019-2022 in the United States. Top row: the number of opioid overdose deaths each year. Subsequent rows show the total age-adjusted death rates per 100,000, then the age-adjusted death rates stratified by sex, race and ethnic origin, specific opioid type involved, or drug co-involvement. Notes: all individuals of Hispanic origin are aggregated, regardless of race (see Methods). The death rate for NHOPI in 2019 is suppressed due to low death counts. In many cases more than one type of opioid is involved. Synthetic Opioids include, for example, drugs such fentanyl (and its derivatives) and tramadol, but excludes methadone. Natural and Semisynthetic Opioids include, for example, oxycodone, hydrocodone, and morphine. Heroin is categorized separately although it is also a semisynthetic opioid. Methadone, a synthetic opioid prescribed to treat opioid use disorder, is categorized separately. Other and Unspecified Narcotics include deaths due to opioid or cocaine-like substances, or mixtures thereof, that were not further sub-categorized on the death certificate. Psychostimulants here refers to psychostimulants with abuse potential, which include methamphetamines but does not include cocaine. See Methods for ICD-10 codes.

|  |  | 2019 | 2020 | 2021 | 2022 |
| --- | --- | --- | --- | --- | --- |
| **Mortality rates** | **All opioid overdose deaths** | 15.5 (49,860) | 21.4 (68,630) | 24.7 (80,411) | 25.0 (81,806) |
| **By gender** | Male | 21.7 (34,635) | 30.4 (48,660) | 34.8 (56,757) | 35.5 (58,385) |
|  | Female | 9.3 (15,225) | 12.3 (19,970) | 14.5 (23,654) | 14.4 (23,421) |
| **By race/ethnicity** | American Indian or Alaska Native | 17.7 (421) | 28.1 (669) | 38.7 (932) | 46.9 (1,109) |
|  | Asian | 1.5 (314) | 2.6 (527) | 2.6 (533) | 3.2 (683) |
|  | Black or African American | 17.3 (7,331) | 26.6 (11,338) | 33.5 (14,537) | 36.6 (15,917) |
|  | Hispanic origin (all races) | 8.8 (5,264) | 13.1 (7,966) | 16.0 (9,921) | 17.3 (10,919) |
|  | More than one race | 7.3 (431) | 12.2 (745) | 15.0 (926) | 16.2 (1,029) |
|  | Native Hawaiian or Other Pacific Islander | NA | 6.0 (39) | 9.7 (62) | 8.3 (55) |
|  | White | 19.2 (35,779) | 25.5 (46,961) | 28.4 (53,022) | 27.6 (51,457) |
| **By opioid type** | Synthetic opioids | 11.4 (36,359) | 17.8 (56,516) | 21.8 (70,601) | 22.7 (73,838) |
|  | Natural and semisynthetic opioids | 3.6 (11,886) | 4.0 (13,471) | 4.0 (13,618) | 3.5 (11,871) |
|  | Heroin | 4.4 (14,019) | 4.1 (13,165) | 2.8 (9,173) | 1.8 (5,871) |
|  | Methadone | 0.8 (2,740) | 1.1 (3,543) | 1.1 (3,678) | 1.0 (3,334) |
|  | Other and unspecified narcotics | 0.5 (1,549) | 0.5 (1,654) | 0.4 (1,452) | 0.3 (1,105) |
| **By drug co-occurrence** | Psychostimulants | 2.8 (8,642) | 4.7 (14,777) | 6.7 (21,371) | 7.2 (23,006) |
|  | Cocaine | 3.8 (11,998) | 4.8 (15,338) | 5.9 (19,250) | 6.6 (21,701) |

We accessed opioid overdose death counts in 5-year age groups for each demographic group (e.g. race/ethnicity + sex; state) to avoid the data suppression that occurs when <10 deaths were observed. To further minimize the impact of this suppression, we also downloaded the *total* number of opioid-related deaths in each group and calculated the “missing” deaths as the difference between the total and the summed age-specific deaths. These missing deaths are then distributed across the age groups with suppressed deaths, using a multinomial distribution with event probabilities set by the national age distribution of opioid-related deaths (refer to *Supplementary Methods*).

We use US period life tables from the National Vital Statistics Systems (NVSS) for the years 2019, 2020, and 2021, and provisional life tables for the year 2022, reported by sex and race/ethnicity, and by state.^20^ Life tables track a population’s mortality experience and are commonly used in demography and actuarial science.^21^

### Life expectancy and years of life lost calculation

We estimate the impact of the opioid crisis on life expectancy in the US by comparing the current life expectancy to an estimate of counterfactual life expectancy in a hypothetical scenario in which no opioid-related deaths are occurring, using a cause-eliminated life table (refer to *Supplementary Methods*). Life expectancy (LE) is the average remaining length of life of individuals at each age, assuming that current age-specific mortality patterns were to remain constant throughout their lifetimes. The reduction in life expectancy due to a cause of death accounts for both age at death and the number of deaths.

The counterfactual life expectancy is extracted from the cause-eliminated life table, which estimates the mortality experience of a population in the hypothetical scenario in which the cause of death in question was completely eliminated from the population.^21^ The cause-eliminated life table is derived from the all-cause life table, using standard demography techniques (refer to *Supplementary Methods*). At each age, the probability of survival is rescaled to account for the absence of this cause of death. Individuals who would have died from this cause of death are still subject to the overall remaining mortality rate.

The difference between the real life expectancies (*rLE*_*age*_) and counterfactual life expectancies (*cLE*_*age*_) reveals the impact of the cause in question on life expectancy:

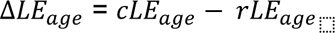

Using this method, we estimate the reduction in life expectancy at birth due to opioid overdose deaths for the entire US population and by demographic group, using opioid-specific and all-cause death counts, stratified by age, from the CDC WONDER database and life tables from the NVSS.

We can calculate years of life lost (YLL) due to a cause of death by multiplying the number of deaths at each age due to this particular cause (*N*_*age*_) by the counterfactual LE in the absence of this cause of death (*cLE*_*age*_). We use the average between the LE at that age and the start of the next age group, since deaths occur throughout the age interval. Total YLL are obtained by summing over all ages:

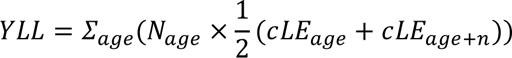

Here, we estimated YLL in the US due to the opioid crisis in 2019, 2020, 2021, and 2022, separately for each demographic group (race/ethnicity and sex, or state).

Previous studies of the YLL from overdose deaths have compared individuals’ age at death to either a specific cutoff age (sometimes alternatively referred to as *potential years of life lost*),^15^ or to the remaining LE for that age.^14,16,17^ In this study, we compare the age at death to our estimated counterfactual remaining LE. We believe this better matches the interpretation of YLL as being relative to a counterfactual scenario where all opioid overdose mortality was prevented, but the mortality experienced due to other causes was constant. For causes of death with a measurable impact on the population’s LE, accounting for the increase in LE if this cause was removed is an important component of estimating YLL.

We report death rates, YLL, and LE without uncertainty throughout the paper. The death counts reported in the CDC WONDER database are real counts of the total number of deaths that occurred across the entire US population (not measured from a sample), and are not reported with any error bars. The NVSS life tables are constructed from the same complete national mortality data and are also not reported with any error bars. Although some additional statistical corrections to deal with missing data and misclassification are applied in creating the life tables, no uncertainty associated with those correction mechanisms is reported. The dominant source of error in this analysis is likely from misclassification of deaths due to difficulty for the certifier (e.g. attending physicians or medical examiner) to determine or decide the cause of death. These types of errors are difficult to quantify, and it is typical for similar analyses of US mortality data to report estimates without error bars.^14,19^

Analyses were conducted in R (version 4.4.0). These results and more can be found online on our interactive web app, which visualizes data on the opioid crisis: https://annehebert.github.io/dashboard.html

## Results

Opioid overdose deaths in the United States have been on the rise nearly every year since the late 1990s and have increased dramatically since 2019 (*Supplementary Figure S*1). From 2019 to 2022 there was a 64% increase in the number of opioid-related deaths, reaching 81,806 deaths (25.0 per 100,000) in 2022 (Table 1). In 2022, men had a 2·5 times higher rate of opioid-related deaths than women. In all race/ethnicity groups, opioid overdose death rates increased substantially since 2019. In 2022, the highest death rates were amongst American Indian/Alaska Native (AI/AN) and Black/African American people, at 46.9 and 36.6 deaths per 100,000, respectively. The recent sharp increase in overdose deaths is primarily driven by synthetic opioids (mainly fentanyl),^22^ whose death rate doubled from 2019 to 2022 (*Supplementary Figure S*1a). Polysubstance use also rose in this period, primarily of cocaine or psychostimulants with abuse potential, which include methamphetamines (from now on referred to as psychostimulants in this text). By 2022, opioid overdose death rates with co-involvement of psychostimulants or cocaine had increased to 7.2 and 6.6 deaths per 100,000, respectively.

In the years 2019-2022, most opioid overdose deaths occurred among young adults (**Figure 1a**). The median age of individuals dying of opioid overdoses was 40 years old for men, and 41 for women in 2022. Almost all ages experienced an increase in the number of deaths from 2019 to 2022. While the number of opioid-related deaths varies between sexes, the percentage of deaths involving opioids is consistently high, exceeding 20% at each age between 22-36 years for women, and between 23-40 years for men (**Figure 1b**). Polysubstance use also occurred across all adult ages. Death rates involving both psychostimulants and opioids were highest among young adults aged 30-40 years, especially among men, whereas cocaine co-involvement death rates were more uniform across adults aged 30-60 years (**Figure 1d**).

**Figure 1:**
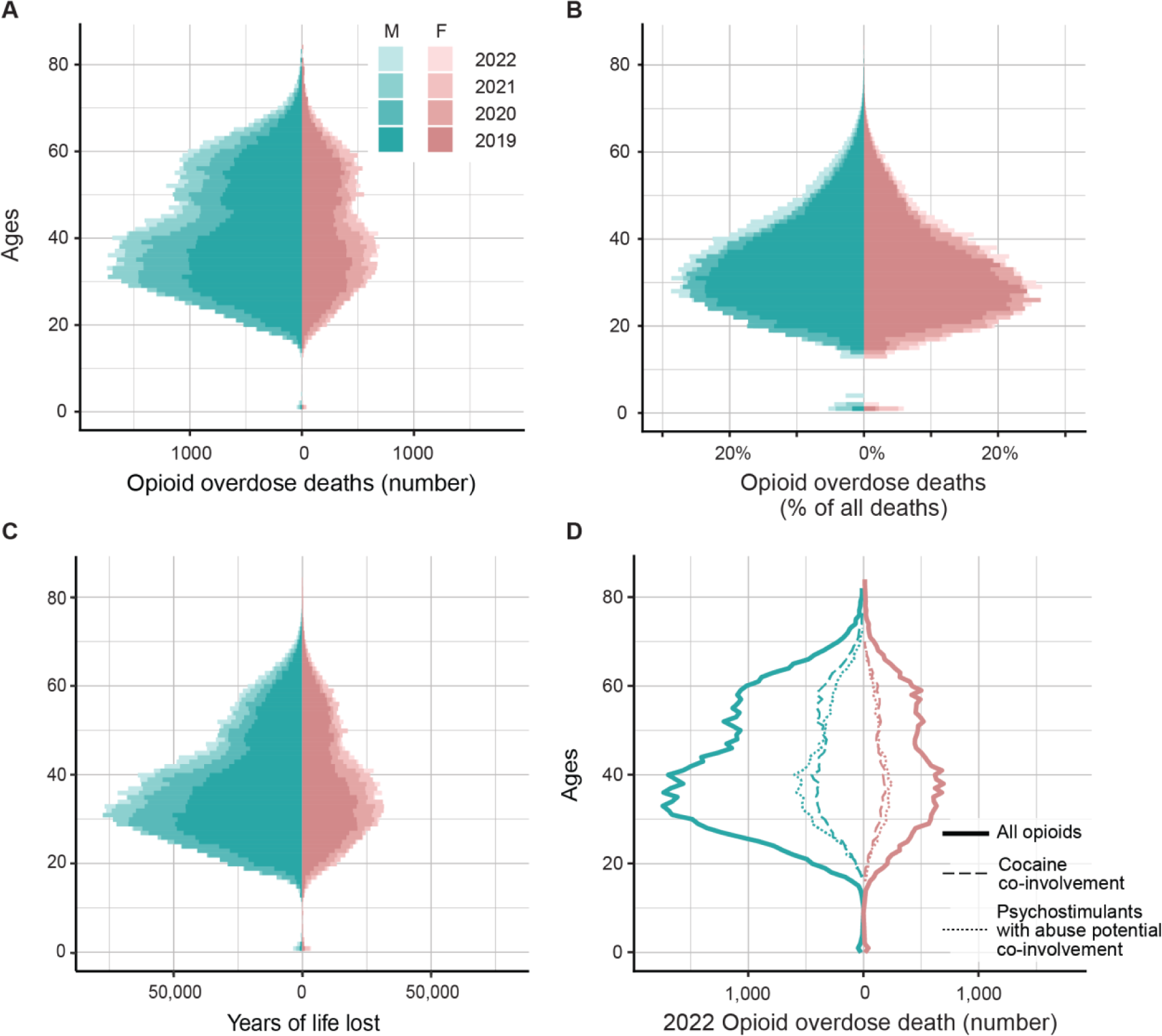
Age distributions of opioid overdose deaths in the United States. **(a)** Age distribution of opioid overdose deaths by sex, in total numbers for 2019 and 2022, and **(b)** the percentage of all deaths that are due to opioid overdoses at each age, by sex, for 2019 and 2022. **(c)** the years of life lost to all opioid overdose deaths at each age, by sex, for 2019 and 2022. **(d)** 2022 age distribution of opioid overdose death rates by sex and polysubstance use.

The life expectancy at birth (LE) of the US population is considerably shortened due to the opioid crisis. We found that LE was reduced by 0·52 years in 2019, and 0·67 years in 2022 (**Table 2**). In 2022, almost half of this loss in LE was due to deaths involving either cocaine or psychostimulants in addition to opioids. Stratifying the analysis by race and sex, we found all groups had a larger loss in LE in 2022 compared to 2019 (**Figure 2a**, *Supplementary Table* 1). In this period, the loss in LE of Asian men, Hispanic men, and AI/AIN men and women more than doubled. In 2022, the LE of AI/AN men was reduced by 1·5 years, followed by Black and white men (1·1 and 0·96 years, respectively). The LE of AI/AN women was reduced by 1 year, followed by white and Black women (0·55 and 0·53 years, respectively).

**Figure 2:**
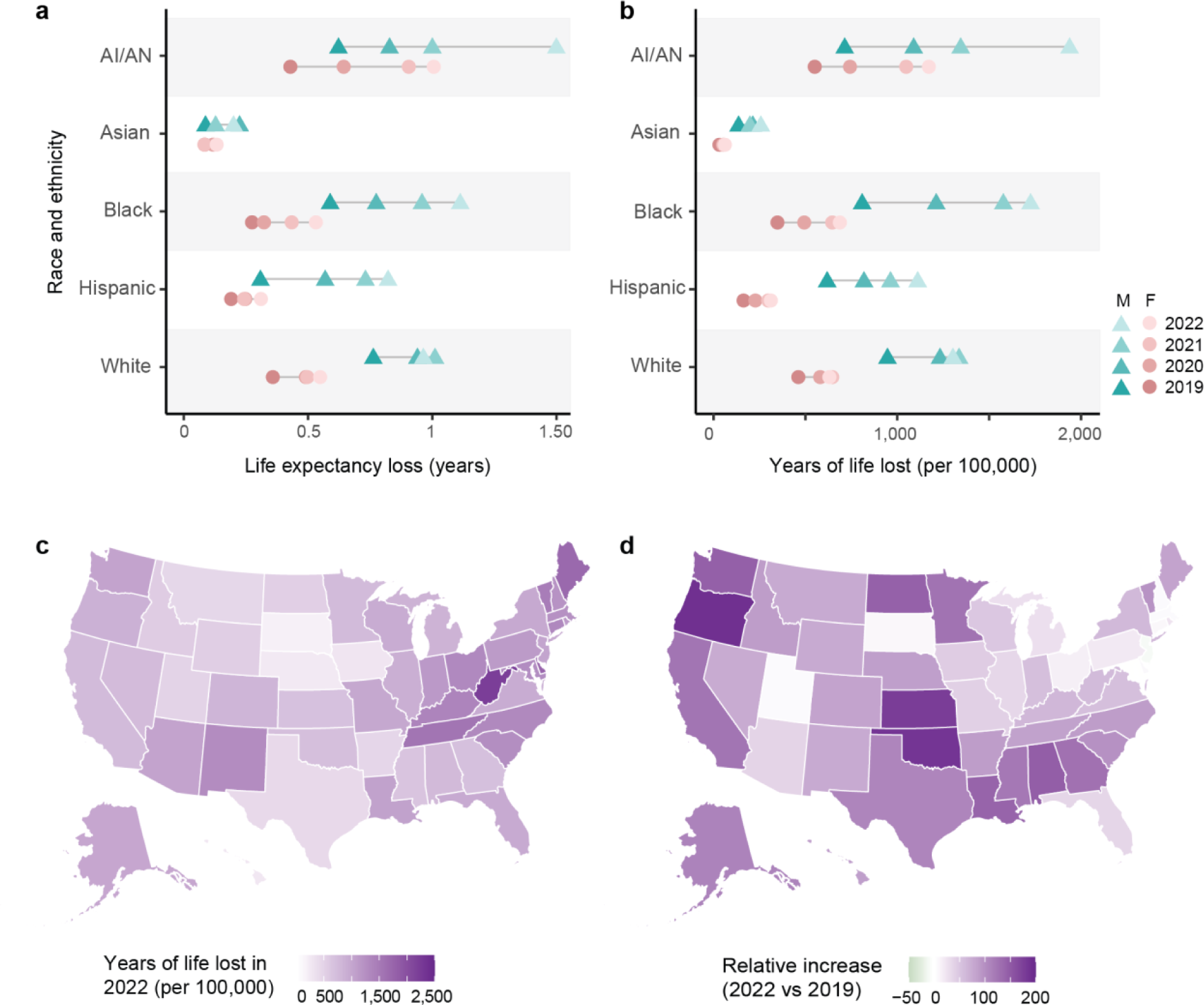
Loss in life expectancy and years of life lost per capita in the US. **(a)** Loss in life expectancy and **(b)** Years of life lost per capita due to opioid-related deaths for each sex and race/ethnicity group, for years 2019-2022. **(c)** Years of life lost per capita due to opioid overdose deaths in 2022, by state, and **(d)** relative increase in years of life lost per capita between 2019 and 2022, by state. Losses in life expectancy are estimated by comparing real-life expectancies to estimated counterfactual life expectancies, describing a scenario in which opioid-related mortality is eliminated. Years of life lost are found by comparing age at death to the counterfactual remaining life expectancy at the age of death.

**Table 2:**
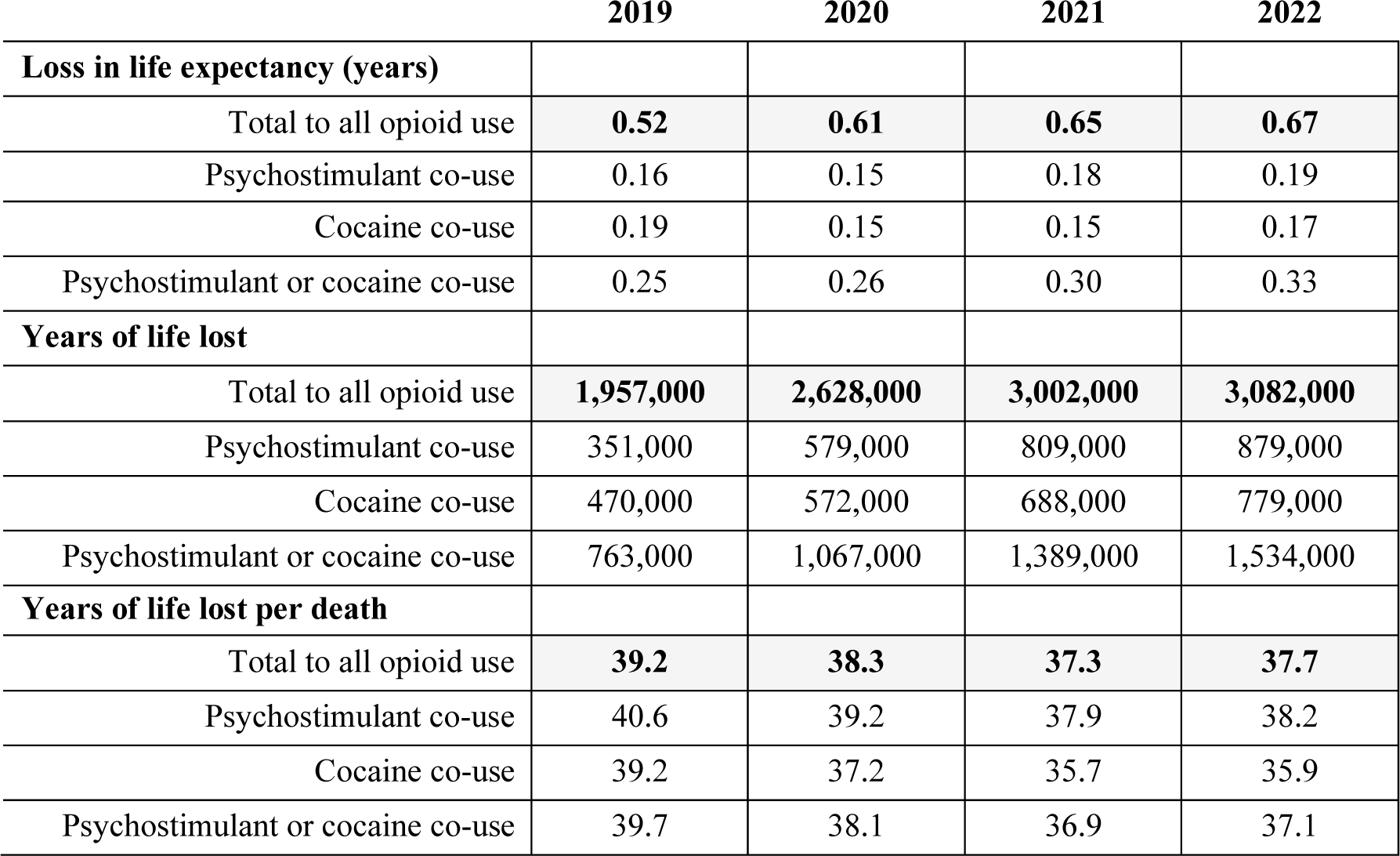
Loss in life expectancy and years of life lost in the US due to opioids. Loss in life expectancy (top), years of life lost (middle), and years of life lost per death (bottom) due to opioid overdose deaths in 2019, 2020, 2021, and 2022. Also shown are the contributions from overdose deaths involving co-use of psychostimulants with abuse potential or cocaine in combination with opioids. Rows reporting “psychostimulant co-use” include any overdose deaths that involve opioids and psychostimulants, which may or may not also involve cocaine. Rows reporting “cocaine co-use” include any overdose deaths that involve opioids and cocaine, which may or may not also involve psychostimulants. Rows reporting “psychostimulant or cocaine co-use” include deaths involving either or both categories of stimulants in addition to opioids. Losses in life expectancy are estimated by comparing real life expectancies to estimated counterfactual life expectancies, describing a scenario in which all opioid-related mortality is eliminated. Years of life lost are found by comparing age at death to the counterfactual remaining life expectancy at age of death.

|  | 2019 | 2020 | 2021 | 2022 |
| --- | --- | --- | --- | --- |
| **Loss in life expectancy (years)** |  |  |  |  |
| Total to all opioid use | **0.52** | **0.61** | **0.65** | **0.67** |
| Psychostimulant co-use | 0.16 | 0.15 | 0.18 | 0.19 |
| Cocaine co-use | 0.19 | 0.15 | 0.15 | 0.17 |
| Psychostimulant or cocaine co-use | 0.25 | 0.26 | 0.30 | 0.33 |
| **Years of life lost** |  |  |  |  |
| Total to all opioid use | **1,957,000** | **2,628,000** | **3,002,000** | **3,082,000** |
| Psychostimulant co-use | 351,000 | 579,000 | 809,000 | 879,000 |
| Cocaine co-use | 470,000 | 572,000 | 688,000 | 779,000 |
| Psychostimulant or cocaine co-use | 763,000 | 1,067,000 | 1,389,000 | 1,534,000 |
| **Years of life lost per death** |  |  |  |  |
| Total to all opioid use | **39.2** | **38.3** | **37.3** | **37.7** |
| Psychostimulant co-use | 40.6 | 39.2 | 37.9 | 38.2 |
| Cocaine co-use | 39.2 | 37.2 | 35.7 | 35.9 |
| Psychostimulant or cocaine co-use | 39.7 | 38.1 | 36.9 | 37.1 |

Years of life lost (YLL) is a measure that incorporates both the number of people dying and the difference between their actual and expected age at death. We estimate that 3·1 million years of life were lost to opioid overdoses in the US in 2022 alone (**Table 2**). In total, 57% (1,125,000) more years of life were lost to opioids in 2022 than 2019. We also found that on average in 2022, 38 years were lost with each opioid-related death. Most of the years of life lost are among younger adults (**Figure 1c**). The YLL per capita varies across race and ethnicity groups, but across groups, men lost more years than women (**Figure 2b**, *Supplementary Table* 1). Amongst women, AI/AN women lost the most years per capita throughout the period (1,070 per 100,000 in 2022). In 2019, white men lost the most years per capita (947 per 100,000), however, by 2022, AI/AN and Black men lost more years per capita (1,940 and 1,730 per 100,000, respectively). Each year, Asian men and women lost the fewest years of life. Psychostimulants with abuse potential or cocaine used in combination with opioids contributed a large portion of the YLL, from 39% in 2019 to 50% in 2022 (**Table 2**). In 2019, cocaine was involved in more loss of life than psychostimulants, but through 2020-2022 psychostimulants were the larger contributor.

There is high geographic variation in the severity of the opioid crisis across the United States. In 2022, 22 states lost more than 1,000 years of life per 100,000. West Virginia saw the most YLL per capita (2,200 years per 100,000), followed by Delaware and Maine (1,800 and 1,700 years per 100,000, respectively) (**Figure 2c**, *Supplementary Table* 3). In contrast, Hawaii, Nebraska, and South Dakota all lost nearly ten times fewer years per capita than West Virginia (280, 260, and 200 YLL per capita). Between 2019 and 2022, the YLL doubled or more in 16 states, 6 of which are in the south (**Figure 2d**). The loss in life expectancy also varies, with 10 states losing more than 1y in 2022 (up to 1·7 years in West Virginia) and 15 states losing between 0·2 -0·5 years (*Supplementary Figure* S2, *Supplementary Table* 3).

Polysubstance use was implicated in close to 50% of all opioid overdose deaths in 2022, a proportion that has been increasing since 2012 (*Supplementary Figure* S1d). However, the contribution of cocaine vs psychostimulants with abuse potential to the overall prevalence of polysubstance involvement differs substantially by demographic group and by state. In 2022, cocaine was reported to be involved in 49% of opioid-related deaths in the District of Columbia vs 0% in South Dakota, while psychostimulant co-involvement varied from 53% in Alaska to 5% in Connecticut (**Figure 3a, b**). In general, states with the highest cocaine co-involvement have the lowest psychostimulant co-involvement, while total polysubstance involvement varied less (between 28-61% in all states and 40-60% in 30/50 states) (*Supplementary Figure* S3). We estimated that overall polysubstance use contributed to about half of the years lost and life expectancy reduction to opioid-related deaths across all racial/ethnic groups (**Figure 3c**, *Supplementary Table* 2). Relative loss of life from psychostimulant co-use was highest amongst AI/AN people (42% and 38% of total YLL among AI/AN women and men, respectively), whereas cocaine co-use was highest among Black people (46% and 42% of YLL among Black women and men, respectively).

**Figure 3:**
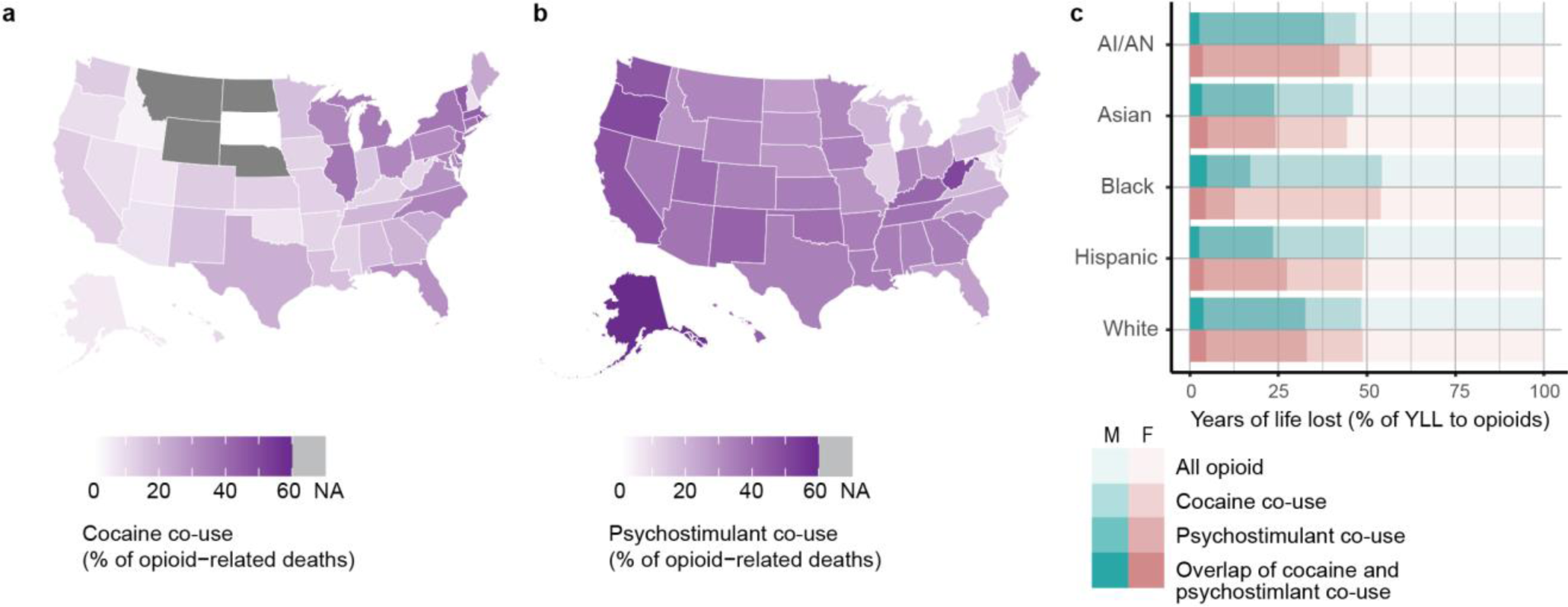
Polysubstance use with opioids in the US in 2022. **(a)** Percentage of opioid-related deaths in 2022 that also involved cocaine, by state. **(b)** Percentage of opioid-related deaths in 2022 that also involved psychostimulants with abuse potential, by state. **(c)** Percentage of the years of life lost to opioids in 2022 for which either cocaine or psychostimulants were also involved, by race/ethnicity and sex group. Psychostimulants here refer to psychostimulants with abuse potential, which include methamphetamines. States with suppressed counts for deaths involving opioids and cocaine (refer to Methods), for which we therefore cannot report the percentage of co-use, are shaded in grey. “Psychostimulant co-use” includes any overdose deaths that involve opioids and psychostimulants, which may or may not also involve cocaine. “Cocaine co-use” includes any overdose deaths that involve opioids and cocaine, which may or may not also involve psychostimulants. “Overlap of cocaine and psychostimulant co-use” includes any deaths involving *both* categories of stimulants in addition to opioids.

## Discussion

The opioid crisis in the United States has been intensifying over the last 25 years, reaching 81,806 opioid overdose deaths in 2022. In this study, we estimate the reduction in life expectancy at birth due to opioid-related mortality, each year from 2019 to 2022. Life expectancy in the US has been stagnating and then declining since 2015, dropping by 2·4 years from 2019-2021.^20^ While this decline is due to a variety of factors, the opioid crisis is likely the largest contributor after COVID-19.^23^ Although there was an increase of 1.1 years from 2021 to 2022, life expectancy still remains lower than in 2004. In this study, we estimate that in 2022 alone, the opioid crisis reduced the US population’s life expectancy (LE) at birth by 0·67 years, compared to 0·52 years in 2019. This surge in fatal opioid overdoses at the onset of the COVID-19 pandemic coincided with disruptions in access to treatment and harm reduction programs, and increased social isolation during lockdown periods.^24^

The years of life lost to opioid overdoses highlight both the large number of deaths and how many are among young people: opioid overdoses are a leading cause of death in young adults nationwide, causing more than 20% of all deaths in 20-39 year olds (**Figure 1b**). We find the years of life lost (YLL) to opioids increased annually, from 2 million in 2019 to 3.1 million in 2022. On average, in 2022, 38 years of life were lost per opioid-related death. While other leading causes of death result in more total YLL, the years lost *per death* are much lower: an estimated 15 years are lost per cancer death, 14 per stroke death, and 9 per COVID-19 death.^25–27^

Using these metrics, we find substantial variations in the burden of opioid-related deaths across demographic groups. In 2019, white men, followed by Black/African American men had the highest burden of opioid overdose deaths. However, by 2022, American Indian/Alaska Native (AI/AN), Black, and white men were all losing the most years of life per capita and experiencing the largest reduction in LE (close to or exceeding 1 year). Amongst women, AI/AN women had the highest burden each year, with a loss in life expectancy of 1 year in 2022. AI/AN, Black, and Hispanic people experienced the largest increases in burden over this time, with years of life lost and loss in life expectancy roughly doubling in 2022 compared to 2019. Our results display a rapidly rising burden of opioid overdose deaths among Black, Hispanic, and Native American peoples and add to previous work finding that contrary to early beliefs and media portrayals, the opioid crisis is no longer simply a “rural white problem”. Now severely affecting people of all races and ethnicities, the opioid crisis will only be slowed if effective, evidence-based treatments are equitably delivered to all.

Our results show a large increase in the metrics of opioid mortality burden in the US, compared to studies in earlier years of the opioid crisis. Previous studies found that in 2016, US life expectancy at age 15 was reduced by 0·36 years, and 1·7 million years of life lost to opioids.^18,19^ Relative to these studies, we estimated that these metrics nearly doubled by 2022, in just six years. A recent study estimated 2·9 million years of life lost in the US in 2021 to unintentional opioid-related deaths alone by comparing age at death to 2019 life expectancy estimates.^18^ In contrast, for each year in our study period, we compared age at death to counterfactual life expectancy estimates for the same year. This allows us to quantify the impact of opioid-related mortality before and during the COVID-19 pandemic, given the backdrop of increases in other causes of death.

Metrics based solely on overdose deaths are not complete measures of burden, as there are many other impacts of this crisis. Individuals suffering from OUD have reduced quality of life and economic participation, and impact the lives of family and community members.^28,29^ OUD is also associated with high mortality rates across major causes of death, such as from infection from bloodborne pathogens such as HIV and HCV, suggesting that the opioid crisis is responsible for additional deaths.^30^ Future work incorporating other metrics of burden such as nonfatal overdose hospitalization rates or elevated mortality rates from other causes may allow for a more complete picture.

This study is based on data extracted from death certificates, which brings some limitations. First, there are guidelines but no standardized process for determining and reporting the cause of death, which leaves room for biases and leads to probable undercounting of opioid overdose deaths.^31^ Secondly, due to misclassification of drug types involved in overdose deaths, there is likely underreporting of opioid overdose deaths, varying by state.^32^ Additionally, racial misclassification, which varies across race/ethnicity groups, may add error to estimates of all-cause and opioid-related mortality, in particular for people of color.^33^ Another source of error in our estimates stems from the data suppression of low death counts (refer to Methods). While we minimize the effects of this suppression by redistributing the missing counts, this adds some uncertainty to our estimates, especially for small groups. Altogether, this leads us to conclude that we are likely underestimating the total YLL to opioid overdose mortality nationally and that our estimates of smaller demographic groups’ opioid-related burden may have larger errors.

Our study focuses on opioid-driven mortality only in the US, which has the estimated highest per capita and absolute burden globally. However, a parallel opioid crisis is unfolding in Canada. ^34–36^ The Public Health Agency of Canada estimates that from 2019 to 2021 there was a 113% increase in the rate of opioid-related deaths (10 to 21·3 per 100,000), compared to a 59% increase in the US over the same period (15·5 to 24·7 per 100,000).^36^ Less appreciated is that many other countries around the world are now experiencing rapidly rising rates of opioid overdose and have recently seen increases in opioid prescribing mirroring that in the US in prior decades.^35,37,38^ The Global Burden of Disease Study 2021 estimated that 36 countries had opioid overdose death rates greater than 1/100,000, concentrated in Europe and Central/Western Asia.^25^ Better quantification of recent changes in the international burden is needed, in addition to urgent policy responses to prevent escalation to levels of the US crisis.

Our study highlights the staggering burden of the US opioid crisis and its acceleration during the COVID-19 pandemic. The US urgently needs comprehensive policy responses that match the scope of this national public health emergency to save hundreds of thousands of lives in the next few years.^35,39^ Our results suggest several areas of focus. Although opioid overdose death counts are themselves high, the disproportionate burden of these deaths on life expectancy and years of life lost is due to the high burden among younger adults. Opioid dependency most often starts during adolescence or young adulthood and policies that specifically focus on preventing and treating addiction in this age group are critical. Two of the populations currently experiencing the largest increases in opioid mortality burden and losing the most life expectancy - Black and AI/AN men - are also the two groups that experience the highest rates of incarceration in the US. Interventions are needed to better treat substance abuse in incarcerated and newly released individuals. While medication-assisted treatment is imperative for the millions of Americans currently living with opioid use disorder and at risk of fatal overdose, the crisis is unlikely to turn around unless the underlying structural drivers of opioid use and so-called “deaths of despair” are addressed. The rapid increase in high-potency fentanyl over the past decade, and the more recent increase in polysubstance overdoses, suggest the important role of the supply chain of illicit drugs in the trajectory of the opioid crisis. Innovative domestic law enforcement and foreign policy approaches are needed to control the massive flow of opioids into the US and their distribution throughout the country. Finally, the acceleration of opioid overdose deaths during the COVID-19 years - after a few promising years of slowdown - demands that pandemic preparedness activities include efforts to avoid the breakdowns in social, economic, and health systems that drive increases in substance abuse. Social isolation, job loss, reduced access to health services, disproportionate levels of severe COVID-19 among Black, AI/AN and Hispanic communities, and supply chain disruptions are all implicated in increased opioid overdose deaths from 2020 onwards. Although major COVID-19 policies included important measures to facilitate OUD treatment, this approach was clearly not sufficient.

## Data sharing statement

All data used in this study is publicly available from the Center for Disease Control and Prevention and the National Center for Health Statistics.

CDC WONDER’s Multiple Cause of Death database: https://wonder.cdc.gov/mcd.html

National Vital Statistics System: https://www.cdc.gov/nchs/nvss/life-expectancy.htm

All analysis code will be available after publication: https://github.com/annehebert/OpioidsYLL

## Data Availability

Our code is open access on GitHub: https://github.com/annehebert/OpioidsYLL
All data produced in the present study are available upon reasonable request to the authors, and are available online at:
https://wonder.cdc.gov/mcd-icd10-expanded.html
https://www.cdc.gov/nchs/nvss/life-expectancy.htm

https://wonder.cdc.gov/mcd-icd10-expanded.html

https://www.cdc.gov/nchs/nvss/life-expectancy.htm

## Acknowledgements

*Funding*: This research did not receive any specific grant from funding agencies in the public, commercial, or not-for-profit sectors.

