## Supplemental material for "Impact of opioid overdoses on US life expectancy and years of life lost, by demographic group and stimulant co-involvement: a mortality data analysis from 2019-2022"

**Supplement**

**Contents:**

**Supplemental Methods:** Counterfactual life expectancy and years of life lost page 2
**Supplemental Methods:** Redistribution of suppressed death counts page 3
**Supplemental Methods:** Data page 3

**Supplement Figure 1:** Trends in opioid overdose deaths in the United States from 1999 to 2022. page 4
**Supplement Figure 2:** US loss of life expectancy by state, 2019-2022. page 5
**Supplement Figure 3:** Opioid and stimulant co-involvement in the US in 2022. page 5

**Supplement Table 1:** Years of life lost in the US due to opioids from 2019 to 2022. page 6
**Supplement Table 2:** Years of life lost and life expectancy reduction to polysubstance use in 2022 in the US, by race and gender. page 7
**Supplement Table 3:** Years of life lost and life expectancy reduction by state, in 2019 and 2022. page 8

**References** page 9

### Supplemental Methods

#### Counterfactual life expectancy and years of life lost

In general, the impact of a particular cause of death on LE can be estimated by finding the difference between the real LE and the counterfactual LE (what the life expectancy would have been in the absence of the cause). The counterfactual life expectancy is derived in the cause-eliminated life table (sometimes referred to as cause-deleted life table), which estimates the mortality experience of a population in the hypothetical scenario in which the cause of death in question were completely eliminated from the population.

We derive the life tables for the scenario in which all opioid-related deaths in the US are eliminated, following the method in the textbook Demography: Measuring and Modeling Population Processes.[1] We start with all-cause life tables from the NVSS (given by race/ethnicity and sex, or state, and by year). We first abridge them into age groups matching the death counts data (<1 years, 1-4 years, and 5-year age groups from 5-100 years), using standard methods.[2] Note that when estimating the years of life lost by sex and year for Figure 1, we used single year age groups, so we don't abridge the NVSS life tables.

We derive the cause-eliminated life table from the all-cause life table, using the fraction of all deaths that are not opioid-related, in each age group:

$$R_{age}^{-opioids} = \frac{D_{age} - D_{age}^{opioids}}{D_{age}}.$$

$D_{age}$ and $D_{age}^{opioids}$ are the number of all-cause and opioid-related deaths in each age group, respectively. These are obtained from CDC WONDER for each year (2019, 2020 and 2021), by 5-year age group and demographic group. However, when estimating years of life lost stratified by sex only, we accessed death counts by single year ages.

With this scaling factor, the cause-eliminated life table values (labeled as *-opioids*) can be calculated from the all-cause life table values, according to the following equations (see Preston, Heuveline and Guillot textbook for more details).

Here *n* refers to the width of the age groups. For the first age group, *n*=1, for the second, *n*=4, and for all subsequent age groups *n*=5. The probability of survival is scaled up to account for removing this cause of death:

$$p_{age}^{-opioids} = (p_{age})^R$$

The number of people surviving at each age is:

$$l_{age+n}^{-opioids} = l_{age}^{-opioids} \times p_{age}^{-opioids}$$

The probability of death within the age interval is:

$$q_{age}^{-opioids} = 1 - p_{age}^{-opioids}$$

And the number of individuals who die within the age interval is:

$$d_{age}^{-opioids} = q_{age}^{-opioids} \times l_{age}^{-opioids}$$

For each age, the difference between the real ($rLE_{age}$) life expectancy from the all-cause life table and the counterfactual ($cLE_{age}$) life expectancy from the cause-eliminated life table reveals the impact of opioid mortality on life expectancy:

$$\Delta LE_{age} = cLE_{age} - rLE_{age}$$

Thus, we estimate the reduction in life expectancy at birth due to opioid overdose deaths for the entire US population and by demographic group, using opioid-specific and all-cause death counts, stratified by age, from the CDC WONDER database and life tables from the NVSS.

We can calculate years of life lost (YLL) due to a cause of death by multiplying the number of deaths at each age due to this particular cause ($N_{age}$) by the remaining counterfactual LE in the absence of this cause of death. Deaths are occurring throughout the age group, rather than all at the start of the age group, so we average between the remaining counterfactual LE at the start of the current age group ($cLE_{age}$) and at the start of the next age group ($cLE_{age+n}$). Total YLL are obtained by summing over all ages:

$$YLL = \Sigma_{age}(N_{age} \times \frac{1}{2}(cLE_{age} + cLE_{age+n}))$$

State life tables:

When estimating the state-specific cause-eliminated life tables, we used the state-specific all-cause life tables from NVSS for the years 2019 and 2020. However, the state-specific life tables are unfortunately not yet available for the years 2021 and 2022, as they are typically released more than 3 years delayed. In order to best approximate the state-specific mortality experiences in 2021 (and 2022), we calculated the change in age-specific probabilities of death (the $q$ values in the life tables) from 2020 to 2021 (and the changes from 2020 to 2022) from the national US population life tables.

$$\Delta q_{age}(2021) = q_{2021,age} - q_{2020,age}$$
$$\Delta q_{age}(2022) = q_{2022,age} - q_{2020,age}$$

This gives us a measure of how much the probability of death at each age changed. We then apply this shift to the 2020 state-specific, age-specific probabilities of death.

$$q_{state,2021,age} = q_{state,2020,age} + \Delta q_{age}(2021)$$
$$q_{state,2022,age} = q_{state,2020,age} + \Delta q_{age}(2022)$$

From these new $q$ values we then re-calculate our new estimate for the 2021 (and 2022) state-specific life tables.

The absolute value of the changes in age-specific probabilities of death between the 2020 and 2021 US population life tables was between 0% and 21% of the 2021 values, with 72/100 ages having a change of <10%. The absolute value of the changes in age-specific probabilities of death between the 2020 and 2022 US population life tables was between 0% and 28% of the 2022 values, with 88/100 ages having a change of <10%.

**Redistribution of suppressed death counts**

In order to minimize the effects of the data suppression that occurs for death counts <10 (see Methods), we also separately download the total number of opioid-related deaths for each group. From this, we calculate how many deaths were suppressed, and redistribute them across the age groups with suppressed death counts, using a multinomial distribution. We set event probabilities according to the national age distribution of opioid overdose deaths.

When calculating our cause-eliminated life tables by race/ethnicity and sex, or by state, we repeat this redistribution of suppressed death counts 100 times. At each iteration, we estimate the age-dependent scaling factor R, based on the resulting redistributed opioid death age distribution and calculate the cause-eliminated life table. From this we find a distribution of values for the years of life lost and the loss of life expectancy. Throughout the text we report the mean of this distribution. The resulting 95% confidence intervals are vanishingly small (varying in width from 0.4% to 0.0002% of the mean).

**Data**

States throughout the US have different protocols for reporting sex on birth certificates, in terms of whether certifiers have the authority to determine the decedent’s sex listed on the death certificate, or whether there are laws protecting decedents’ gender identity and thus requiring that their self-identified gender be included on the death certificate. Throughout, we use the term 'sex,' in agreement with the National Vital Statistics System's terminology in analyzing the same death certificates data.

**Supplemental Figures**

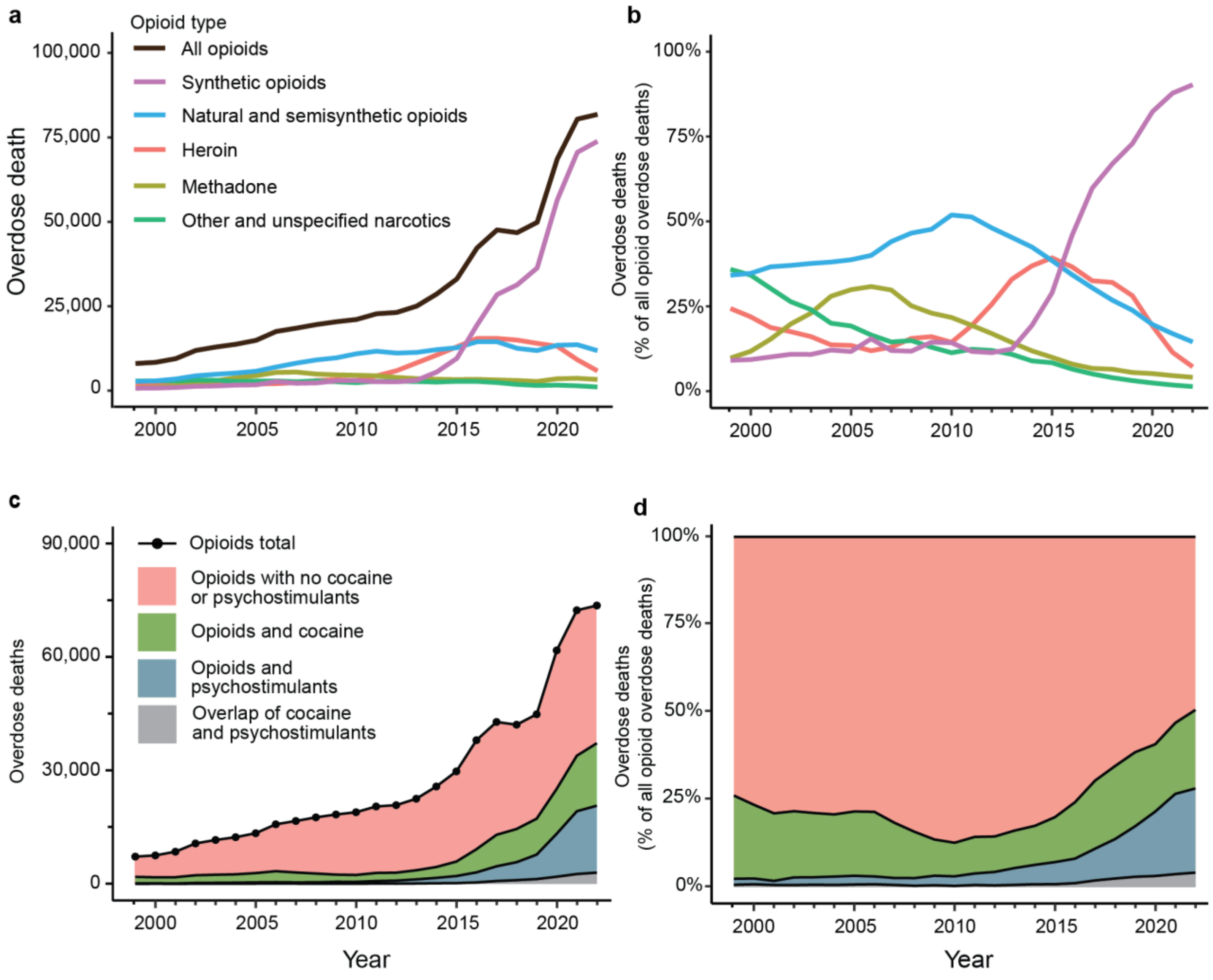


**Supplement Figure 1: Trends in opioid overdose deaths in the United States from 1999 to 2022.** A) Total number of opioid overdose deaths each year, also showing the number of deaths by opioid type. B) Percentage of yearly opioid overdose deaths by opioid type. C) Total number of opioid overdose deaths each year, showing the subset of deaths also involving either cocaine or psychostimulants with abuse potential (e.g. methamphetamines). D) Percentage of yearly opioid overdose deaths by stimulant co-use. Note that some deaths involve more than one type of opioid. Synthetic Opioids include, for example, drugs such fentanyl (and its derivatives) and tramadol, but excludes methadone. Natural and Semisynthetic Opioids include, for example, oxycodone, hydrocodone, and morphine. Heroin is categorized separately although it is also a semisynthetic opioid. Methadone, a synthetic opioid prescribed to treat opioid use disorder, is categorized separately. Other and Unspecified Narcotics include deaths due to opioid or cocaine-like substances, or mixtures thereof, that were not further sub-categorized on the death certificate. Data shown here are death counts from the CDC WONDER Multiple Cause of Death database (see Methods for ICD codes used).

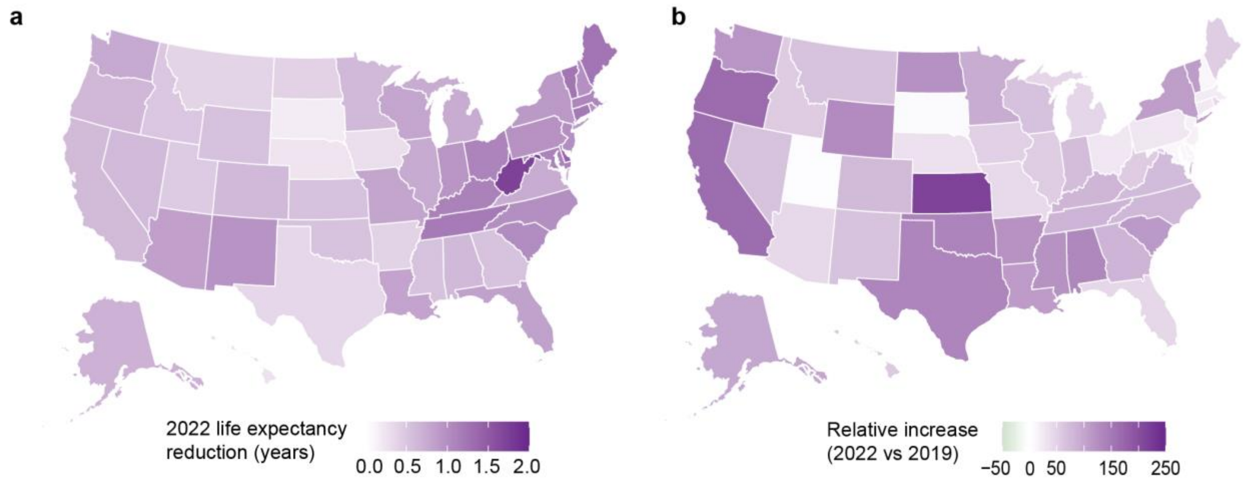


**Supplement Figure 2: US loss of life expectancy by state, 2019-2022.** A) Life expectancy reduction to opioid overdose deaths in 2022, by state and B) relative increase in loss of life expectancy between 2019 and 2022, by state. Life expectancy reductions are estimated by comparing real life expectancies to estimated counterfactual life expectancies, describing a scenario in which opioid-related mortality is eliminated.

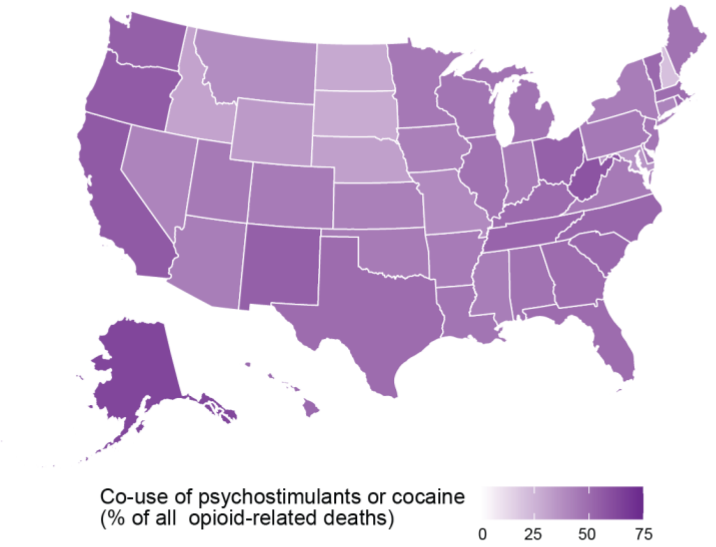


**Supplement Figure 3: Opioid and stimulant co-involvement in the US.** Percentage of opioid-related deaths involving either psychostimulants with abuse potential or cocaine in 2022, by state.

**Supplemental Tables**

| | Sex | **AI/AN** | **Asian** | **Black** | **Hispanic** | **White** |
|---|---|---|---|---|---|---|
| 2022 Loss in Life Expectancy | F | 1.0 | 0.13 | 0.53 | 0.31 | 0.55 |
| | M | 1.5 | 0.20 | 1.1 | 0.82 | 0.96 |
| 2021 Loss in Life Expectancy | F | 0.90 | 0.083 | 0.43 | 0.24 | 0.50 |
| | M | 1.0 | 0.13 | 0.96 | 0.73 | 1.0 |
| 2020 Loss in Life Expectancy | F | 0.63 | 0.12 | 0.32 | 0.25 | 0.49 |
| | M | 0.83 | 0.22 | 0.78 | 0.57 | 0.94 |
| 2019 Loss in Life Expectancy | F | 0.43 | 0.084 | 0.27 | 0.19 | 0.36 |
| | M | 0.62 | 0.087 | 0.59 | 0.31 | 0.76 |
| 2022 Years of Life Lost | F | 14,300 | 6,500 | 150,200 | 97,900 | 622,300 |
| | M | 23,200 | 25,100 | 349,000 | 358,600 | 1,271,000 |
| 2021 Years of Life Lost | F | 13,000 | 5,700 | 140,400 | 92,000 | 640,200 |
| | M | 16,300 | 18,700 | 303,800 | 305,500 | 1,305,700 |
| 2020 Years of Life Lost | F | 9,200 | 5,200 | 106,700 | 69,200 | 578,900 |
| | M | 13,000 | 19,800 | 240,100 | 253,200 | 1,196,300 |
| 2019 Years of Life Lost | F | 6,800 | 3,200 | 74,600 | 48,900 | 461,500 |
| | M | 8,600 | 12,300 | 159,200 | 189,000 | 921,200 |
| 2022 Years of Life Lost per capita | F | 1,170 | 61.7 | 688 | 312 | 630 |
| | M | 1,940 | 257 | 1,730 | 1,110 | 1,300 |
| 2021 Years of Life Lost per capita | F | 1,050 | 55.8 | 646 | 297 | 646 |
| | M | 1,350 | 198 | 1,510 | 963 | 1,340 |
| 2020 Years of Life Lost per capita | F | 742 | 51.1 | 494 | 227 | 580 |
| | M | 1,090 | 214 | 1,210 | 819 | 1,230 |
| 2019 Years of Life Lost per capita | F | 551 | 31.9 | 348 | 163 | 461 |
| | M | 714 | 136 | 809 | 618 | 947 |

**Supplement Table 1: Years of life lost in the US due to opioids from 2019 to 2022.** Years of life lost (top) and loss of life expectancy (bottom) due to opioid overdose deaths in 2019, 2020, 2021, and 2022, by race and ethnicity, and by sex. We only report the years of life lost and the reduction in life expectancy for the 5 largest race and ethnicity groups. There was not enough data from the Native Hawaiian and Other Pacific Islanders group to estimate the years of life lost or the reduction in life expectancy (see Methods).

| | Sex | **AI/AN** | **Asian** | **Black** | **Hispanic** | **White** |
|---|---|---|---|---|---|---|
| Loss in Life Expectancy to psychostimulant co-use | F | 0.46 | 0.10 | 0.11 | 0.14 | 0.22 |
| | M | 0.61 | 0.064 | 0.18 | 0.24 | 0.32 |
| Loss in Life Expectancy to cocaine co-use | F | 0.18 | 0.10 | 0.27 | 0.14 | 0.16 |
| | M | 0.23 | 0.067 | 0.47 | 0.28 | 0.19 |
| Loss in Life Expectancy to both psychostimulant and cocaine co-use | F | 0.55 | 0.11 | 0.31 | 0.19 | 0.30 |
| | M | 0.75 | 0.10 | 0.60 | 0.44 | 0.47 |
| Years of life lost to psychostimulant co-use | F | 6,100 | 1,600 | 18,900 | 26,800 | 205,500 |
| | M | 8,800 | 6,000 | 59,600 | 84,300 | 414,800 |
| Years of life lost to cocaine co-use | F | 1,800 | 1,600 | 68,700 | 24,700 | 126,800 |
| | M | 2,700 | 6,500 | 146,400 | 102,000 | 249,400 |
| Years of life lost to both psychostimulant and cocaine co-use | F | 7,400 | 2,900 | 80,900 | 47,700 | 303,900 |
| | M | 10,900 | 11,600 | 189,300 | 176,700 | 615,300 |
| Years of life lost per capita to psychostimulant co-use | F | 495 | 15.0 | 86.5 | 85.5 | 208 |
| | M | 736 | 61.3 | 295 | 261 | 425 |
| Years of life lost per capita to cocaine co-use | F | 149 | 15.6 | 315 | 78.7 | 128 |
| | M | 225 | 66.2 | 724 | 316 | 256 |
| Years of life lost per capita to both psychostimulant and cocaine co-use | F | 602 | 27.4 | 370 | 152 | 308 |
| | M | 909 | 119 | 936 | 547 | 631 |

**Supplement Table 2: Years of life lost and life expectancy reduction to polysubstance use in 2022 in the US.** Loss in life expectancy (top), Years of life lost (middle), and Years of life lost per capita (bottom) due to co-involvement of opioids and stimulants in 2022, by race/ethnicity and sex.

| State | 2019 Years of Life Lost | 2022 Years of Life Lost | 2019 Years of Life Lost per 100,000 | 2022 Years of Life Lost per 100,000 | 2019 Loss in Life Expectancy | 2022 Loss in Life Expectancy |
|---|---|---|---|---|---|---|
| Alabama | 15,300 | 39,000 | 311 | 768 | 0.265 | 0.587 |
| Alaska | 3,400 | 7,200 | 465 | 985 | 0.348 | 0.641 |
| Arizona | 54,000 | 75,400 | 742 | 1,020 | 0.594 | 0.792 |
| Arkansas | 7,400 | 14,000 | 244 | 460 | 0.181 | 0.377 |
| California | 131,200 | 291,900 | 332 | 748 | 0.234 | 0.558 |
| Colorado | 26,200 | 48,400 | 455 | 828 | 0.352 | 0.573 |
| Connecticut | 43,300 | 48,400 | 1,210 | 1,330 | 0.963 | 1.08 |
| Delaware | 15,800 | 17,800 | 1,620 | 1,750 | 1.23 | 1.30 |
| D.C. | 7,700 | 9,900 | 1,090 | 1,480 | 0.855 | 1.20 |
| Florida | 149,700 | 211,800 | 697 | 952 | 0.574 | 0.771 |
| Georgia | 32,200 | 75,400 | 303 | 691 | 0.289 | 0.492 |
| Hawaii | 2,100 | 4,000 | 150 | 279 | 0.164 | 0.235 |
| Idaho | 5,400 | 11,100 | 303 | 570 | 0.315 | 0.444 |
| Illinois | 83,200 | 112,400 | 657 | 894 | 0.487 | 0.692 |
| Indiana | 49,200 | 778,000 | 730 | 1,140 | 0.541 | 0.876 |
| Iowa | 6,300 | 9,300 | 200 | 290 | 0.192 | 0.264 |
| Kansas | 7,200 | 20,100 | 246 | 686 | 0.171 | 0.499 |
| Kentucky | 37,500 | 62,400 | 840 | 1,380 | 0.617 | 1.07 |
| Louisiana | 20,200 | 48,200 | 435 | 1,050 | 0.379 | 0.764 |
| Maine | 12,700 | 23,700 | 945 | 1,710 | 0.799 | 1.13 |
| Maryland | 78,900 | 789,000 | 1,310 | 1,280 | 1.01 | 1.04 |
| Massachusetts | 79,800 | 86,000 | 1,160 | 1,2320 | 0.889 | 0.961 |
| Michigan | 66,900 | 85,700 | 670 | 854 | 0.512 | 0.690 |
| Minnesota | 18,500 | 42,600 | 328 | 745 | 0.327 | 0.560 |
| Mississippi | 8,600 | 18,800 | 288 | 639 | 0.243 | 0.512 |
| Missouri | 42,500 | 60,700 | 692 | 983 | 0.568 | 0.760 |
| Montana | 2,800 | 5,100 | 258 | 453 | 0.232 | 0.357 |
| Nebraska | 2,700 | 5,000 | 137 | 257 | 0.185 | 0.224 |
| Nevada | 13,300 | 24,100 | 432 | 757 | 0.359 | 0.552 |
| New Hampshire | 15,400 | 16,800 | 1,130 | 1,200 | 0.883 | 0.888 |
| New Jersey | 99,000 | 93,800 | 1,110 | 1,010 | 0.847 | 0.912 |
| New Mexico | 15,200 | 27,000 | 726 | 1,280 | 0.582 | 0.915 |
| New York | 115,700 | 190,100 | 595 | 966 | 0.432 | 0.812 |
| North Carolina | 73,700 | 141,600 | 703 | 1,320 | 0.556 | 0.927 |
| North Dakota | 1,700 | 4,200 | 221 | 537 | 0.188 | 0.370 |
| Ohio | 132,700 | 154,500 | 1,130 | 1,310 | 0.877 | 1.04 |

| | | | | | | |
|---|---|---|---|---|---|---|
| Oklahoma | 9,700 | 28,300 | 246 | 704 | 0.218 | 0.488 |
| Oregon | 12,700 | 37,200 | 301 | 878 | 0.238 | 0.567 |
| Pennsylvania | 119,700 | 145,300 | 935 | 1,120 | 0.764 | 0.896 |
| Rhode Island | 9,600 | 12,300 | 903 | 1,130 | 0.673 | 0.871 |
| South Carolina | 32,800 | 66,700 | 637 | 1,260 | 0.481 | 0.963 |
| South Dakota | 1,600 | 1,800 | 180 | 196 | 0.162 | 0.167 |
| Tennessee | 58,000 | 110,200 | 850 | 1,560 | 0.647 | 1.12 |
| Texas | 58,400 | 127,000 | 202 | 423 | 0.149 | 0.323 |
| Utah | 15,200 | 16,700 | 473 | 495 | 0.446 | 0.416 |
| Vermont | 4,700 | 9,500 | 747 | 1,470 | 0.591 | 1.12 |
| Virginia | 50,900 | 80,900 | 596 | 932 | 0.432 | 0.679 |
| Washington | 32,200 | 80,800 | 423 | 1,040 | 0.352 | 0.676 |
| West Virginia | 24,800 | 39,500 | 1,390 | 2,220 | 1.11 | 1.66 |
| Wisconsin | 36,500 | 55,000 | 626 | 934 | 0.474 | 0.725 |
| Wyoming | 1,800 | 3,200 | 309 | 544 | 0.235 | 0.506 |

**Supplement Table 3: Burden of opioid mortality by state in 2019 and 2022**. Years of life lost, years of life lost per 100,000 and loss in life expectancy (years) to opioid related deaths in 2019 and 2022, by US state.
